# Lower mitochondrial DNA abundance in blood cells is associated with higher general morbidity and all-cause mortality: a 30-year prospective epidemiological study

**DOI:** 10.64898/2026.02.10.26345983

**Authors:** Attila A. Sebe, Juulia H. Lautaoja-Kivipelto, Jari Jokelainen, Juho T. Väänänen, Sini Skarp, Karri Parkkila, Risto Kerkelä, Eija Pirinen, Olavi Ukkola

## Abstract

**Background:** Low mitochondrial DNA (mtDNA) abundance in blood cells has been associated with various diseases and major causes of mortality. However, the causal link and molecular mechanisms involved remain unclear, and previous studies have had limited follow-up durations. To address these gaps, we examined the relationship of blood mtDNA abundance with all-cause and cause-specific mortality over three decades, and integrated blood transcriptomic profiling to explore underlying molecular mechanisms. Our goal was to improve the understanding of blood mtDNA abundance as a potential early biomarker for major clinical conditions.

**Methods and findings:** We utilized the clinical and epidemiological data from the prospective OPERA cohort (Oulu Project Elucidating Risk of Atherosclerosis), comprising 1045 individuals initially assessed in the 1990s and followed for over three decades, with a second visit in the 2010s. Blood mtDNA was quantified using real-time quantitative polymerase chain reaction at both time points, and RNA-sequencing was performed on 450 follow-up blood samples. Lower blood mtDNA levels in the 1990s samples were significantly associated with increased overall morbidity and all-cause mortality, assessed up to the end of 2022. Similar trends were observed in a subset of 597 participants from the 2010s. When causes of death were categorized as “cardiovascular”, “cancer”, or “other”, lower blood mtDNA levels predicted higher mortality across all categories. Transcriptomic analysis of the follow-up samples suggested that blood mtDNA variation may be linked to subclinical inflammation involving innate immunity.

**Conclusions:** Blood cell mtDNA abundance shows promise as an early biomarker for general morbidity and mortality in the middle-aged population, although it is not specific to distinct causes of death. The underlying pathomechanism of lower blood mtDNA levels may involve inflammatory processes. These findings, combined with the three-decade follow-up, support the potential use of blood mtDNA in the primary prevention of morbidity and mortality of various etiology.

## 1. Introduction

Mitochondria are essential cellular organelles that, over recent decades, have been extensively studied related to aging and disease. These organelles are well known for their role in ATP production through oxidative phosphorylation (OXPHOS) in all eukaryotic cells [1]. However, they are involved in various other functions, such as the production of reactive oxygen species, biosynthesis of metabolites, cellular signaling, regulation of apoptosis, adaptations to cellular stress and immune responses [2]. To support these diverse functions, mitochondria contain their own circular genome, the mitochondrial DNA (mtDNA), present in multiple copies in each mitochondrion. This genome encodes 13 essential proteins of the OXPHOS system, along with 22 transfer and 2 ribosomal RNAs and these protein-coding genes have no nuclear-encoded equivalents [3]. Mitochondrial content within cells and the amount of mtDNA copies within mitochondria vary widely across different cell types and metabolic conditions in humans [4]. It is commonly assumed that higher mtDNA amount is associated with increased mitochondrial number and function, while lower mtDNA amount is linked to mitochondrial loss and dysfunction, and cellular stress [5]. Therefore, mtDNA amount is often considered as an indicator of mitochondrial health.

Currently, the quantification of mtDNA is one of the most common and affordable methods to assess mitochondrial content and function. Blood is the most accessible sample source for this purpose in humans, even though evidence linking blood mtDNA levels to tissue-specific mtDNA levels and OXPHOS capacity remains limited [6]. Several recent publications have reported associations of blood mtDNA with inflammatory markers [7–11] suggesting that blood mtDNA levels may also reflect systemic inflammation and overall pathological conditions in the body. It is important to note that blood mtDNA abundance usually refers to the amount measured from the cellular components of blood, primarily white blood cells. It is fundamental to recognize that this intracellular mtDNA amount has clearly different implications than the extracellular mtDNA amount measured from cell-free plasma [12,13]. Altogether, the assessment of mtDNA in peripheral blood provides a valuable tool for exploring associations between mitochondrial measures and various clinical and epidemiological variables.

Cardiovascular diseases (CVD) are the leading cause of death worldwide [14,15]. Therefore, it is crucial to initiate prevention of CVD as early as possible in the current progressively aging population. CVD are frequently characterized by mitochondrial dysfunction [16]. Reports have noted the associations of lower blood mtDNA abundance with the higher prevalence of coronary [17–20] and peripheral artery disease [21], and the higher risk of developing acute CVD events [21–24]. CVD risk factors like obesity [7,25], type 2 diabetes [7,22], dyslipidemia [7,22], hypertension [7,17,22] and smoking [26,27] have also been found to be more prevalent with lower blood mtDNA levels. However, some studies reported the lack of such associations [20,26,28,29], especially concerning causality, highlighting the need for further research.

Lower blood mtDNA abundance has been linked to higher CVD mortality [30]. Similar associations have been observed with all-cause mortality in general populations [31–34], and were recently confirmed by a comprehensive meta-analysis combining multiple studies [35]. These relationships have also been reported in patients with peripheral artery [21] or chronic kidney disease [36]. However, publications related to cancer have documented inconsistent results [21,34,37]. The longest follow-up times across individual studies have been approximately 17 years [35]. Although the data about blood mtDNA levels is extensive, the heterogeneity of the reported associations raises questions about the specificity of blood mtDNA as a biomarker. Additionally, currently available evidence does not clarify causality, leaving it uncertain whether low blood mtDNA levels contribute to the disease progression or reflect certain underlying pathological processes. Thus, these findings underscore the need for further longitudinal studies to better understand the prognostic potential of blood mtDNA abundance.

In this study, we aimed to deepen the understanding of blood mtDNA abundance as a predictive biomarker for CVD, as well as all-cause and cause-specific mortality. Utilizing a prospective cohort design with an extended follow-up period of 30 years, we investigated the relationship between blood mtDNA levels, and morbidity and mortality outcomes. To comprehensively evaluate the specificity of blood mtDNA to these outcomes, we expanded our analysis to include a broad spectrum of causes of death beyond CVD. Our findings demonstrate that reduced blood mtDNA levels serve as a significant predictor of both all-cause and cause-specific mortality across multiple outcomes, with predictive capacity extending up to three decades. Furthermore, blood cell transcriptional profiling revealed a strong association between mtDNA abundance and neutrophil degranulation pathway, suggesting a dynamic interplay between inflammation and mitochondria, and highlighting their combined contributions to disease outcomes and mortality risk. Collectively, these results support the utility of blood mtDNA quantification as a valuable tool for morbidity and mortality risk assessment in primary healthcare settings, also highlighting its potential to facilitate early intervention strategies in middle-aged populations.

## 2. Methods

### 2.1. Study design

#### 2.1.1. Baseline

Our research was based on the OPERA (Oulu Project Elucidating Risk of Atherosclerosis) project, a longitudinal epidemiological study initiated in Oulu, Finland in 1991. In the first phase of this project, subjects aged 40-62 years (*n*=1045) with hypertension (*n*=519) and their age- and sex-matched controls (*n*=526) were recruited between 1991 and 1993. At the time of the hospital visit, whole blood samples were collected and stored at the Oulu University Hospital. Selection criteria and baseline information of the study cohorts have been described in detail elsewhere [38], including the most important anthropometric, lifestyle and clinical parameters, and comprehensive laboratory results, all of which were used in the present analyses. None of the subjects included in the study had a known history of cancer.

Data on mortality causes were obtained from the Finnish Causes-of-Death Register. At the time of this study, all-cause mortality data was available up to the end of 2022 and cause of death statistics up to the end of 2021. Diagnoses were based on the International Classification of Diseases and Related Health Problems 9^th^ (ICD-9) and 10^th^ (ICD-10) Revisions. In our study, we defined CVD as the diagnosis of either coronary heart disease or stroke (excluding subarachnoid hemorrhage), or both. Cancer diagnosis included all types of cancer. Causes of death were recorded, when they were listed as the underlying cause of death, immediate cause of death or as the first to third contributing causes of death [39]. When both CVD and cancer diagnoses were present in a subject’s registry, we selected the condition identified as the primary underlying cause of death.

For a comprehensive cardiovascular perspective, we conducted a separate assessment of CVD events, which included both events leading to death and hospitalization. Data on hospitalization events were obtained from the Hospital Discharge Register and were available up to the end of 2014. CVD events included a major coronary heart disease event and stroke (excluding subarachnoid hemorrhage), whichever of them occurred first. In addition to the respective ICD-10 diagnoses, CVD events were registered if a subject had undergone coronary artery bypass graft surgery or angioplasty [39].

Of the original 1045 whole blood samples, 1023 were available for DNA isolation and quantification at the time of our analysis. We divided our entire baseline cohort of 1023 subjects into tertiles based on the amount of blood mtDNA in their blood samples to reveal correlations with mortality data and cardiovascular morbidity while maximizing the strength of the statistical comparison and enabling effective visualization of results. The quantification of blood mtDNA is described in Section 2.2. To account for the potential impact of sex on mtDNA levels [7,22,26,27,32,33], we first created tertiles of low, moderate and high blood mtDNA abundance in the male and female subgroups separately. Subsequently, we pooled the sex-specific tertiles of low, moderate and high blood mtDNA abundance together, respectively. As a result, our final sex-matched tertiles consisted of approximately equal proportion of male and female subjects, with the low, moderate and high blood mtDNA tertiles consisting of 348, 345 and 330 participants, respectively.

#### 2.1.2. Follow-up

In the second phase of the OPERA study, conducted in 2013-2014, 600 of the 813 survivors participated in a second visit at the clinics, including 290 from the group with hypertension and 310 from the controls. The interview and clinical examination were documented by a hospital nurse as during the baseline visit and the results can be found in the respective publication [40]. Mortality data were available from the Finnish Causes-of-Death Register, as described in the baseline section. During the follow-up visit, blood samples were collected followed by buffy coat isolation. Buffy coat samples were stored at the Oulu University Hospital. Of the original 600 samples 597 were available for DNA and 450 for RNA analysis at the time of our study.

Similarly to the baseline analysis, we created sex-matched tertiles of low, moderate and high blood mtDNA abundance of the 597 available samples by pooling together the male and female sex-specific tertiles of low, moderate and high blood mtDNA abundance. The resulting sex-matched tertiles consisted of 221, 183 and 193 subjects, respectively. It should be noted that the tertiles of low, moderate and high blood mtDNA amount of the follow-up cohort are, by definition, not a fraction of the respective baseline tertiles, even though there may be partial overlap regarding the subjects they consist of.

### 2.2. Quantification of mitochondrial DNA

We determined the abundance of mtDNA in blood cells relative to genomic DNA (often referred to as “mitochondrial copy number”, although more precisely, it refers to a relative measure) using real-time quantitative polymerase chain reaction (RT-qPCR) from whole blood samples from the 1990s (*n*=1023) and buffy coat samples from the 2010s (*n*=597).

Independent of the sample type, total DNA extraction was conducted using the salting out method [41]. Briefly, in the 1990s, the collected whole blood samples were stored at −80°C until use. In the 2010s, whole blood samples were collected into EDTA-containing tubes, and from the anticoagulated blood, buffy coats of nucleated cells were stored at −80°C until use. Note that the buffy coat isolation in the 2010s was carried out with centrifugation, not the other commonly used Ficoll-method, and it resulted in a buffy coat containing all white blood cells, including mononuclear and polymorphonuclear cells. From this step onwards, DNA extraction from the whole blood and buffy coat samples was similar [41]. Briefly, both whole blood and buffy coat samples were mixed with buffer A containing 320 mM D-(+)-sucrose (84100, Fluka, Buchs, Switzerland), 10 mM Tris (pH 7.5, T1378, Sigma, Burlington, MA, USA) and 5 mM MgCl_2_ (1.05833.0250, Merck, Darmstadt, Germany) followed by lysis using a buffer B containing 10 mM Tris (pH 8.0, T1378, Sigma), 90 mM NaCl (0278, J.T.Baker, Phillipsburg, NJ, USA) and 9 mM EDTA (1.08418, Merck). After a 15-minute (min) incubation at room temperature (RT), samples were centrifuged for 15 min at 1500 x g at 4°C. The obtained pellets were dissolved in buffer A followed by a centrifugation for 15 min at 1500 x g at 4°C. Supernatant was removed and buffer B and 20% SDS (L4390-500g, Sigma) were added. The samples were further lysed with proteinase K (P6556-1g, Roche, Basel, Switzerland) administration and an overnight incubation at 37°C. The next day, 5 M NaCl was added, and samples were centrifuged for 15 min at 2100 x g at 4°C. Supernatant was collected and DNA was precipitated by absolute ethanol administration. After removal of ethanol, the precipitated DNA is dissolved into TE buffer (10 mM Tris and 0.1 mM EDTA, pH 7.5) followed by DNA measurement using NanoDrop ND-1000 spectrophotometer (Thermo Fisher Scientific, Waltham, MA, USA). Note that due to the differences in initial sample processing, the whole blood samples contain both cellular and cell-free mtDNA, while the buffy coat samples contain primarily mtDNA from the white blood cells. However, because the amount of cell-free mtDNA in whole blood is small in the absence of acute disease [13], our whole blood results mainly reflect the differences in the cellular mtDNA amount.

Next, the DNA samples (2 ng of template per well) were directed to RT-qPCR conducted according to the manufacturer’s protocol with iQ SYBR Green Supermix (1725006, Bio-Rad Laboratories Inc., Hercules, CA, USA). Data analysis was performed using the CFX96 Real-Time PCR Detection System with CFX Maestro software (Bio-Rad Laboratories Inc.) and the efficiency-corrected ΔΔCt method. Primer sequences were as follows: cytochrome b (*CYTB*) were 5′-GCCTGCCTGATCCTCCAAAT-3′ and 5′-AAGGTAGCGGATGATTCAGCC-3′ for mitochondrial and amyloid beta precursor protein (*APP*) were 5′-TGTGTGCTCTCCCAGGTCTA-3′and 5′-CAGTTCTGGATGGTCACTGG-3 for genomic DNA analysis. Data were normalized to an internal control included in each run to minimize the run-to-run variation among the groups before combining all data for the final analysis.

### 2.3. RNA extraction

Total RNA was isolated from buffy coat samples collected during follow-up using the PAXgene Blood RNA Kit (762174, Qiagen, Hilden, Germany) according to the manufacturer’s instructions. Briefly, buffy coat samples were lysed in PAXgene Blood RNA tubes, centrifuged for 10 min at 3270 x g at RT, and pellets were dissolved in RNase-free water. Next, after a 10-min centrifugation at 3270 x g at RT, pellets were dissolved in resuspension buffer together with binding buffer and proteinase K. Samples were incubated for 10 min at 55°C on a shaker. Next, samples were transferred into a PAXgene Shredder spin column, centrifuged for 3 min at 16 000 x g and pellets were washed with ethanol. Once transferred into PAXgene RNA spin columns, samples were centrifuged for 1 min at 14 000 x g at RT and washed with wash buffer. Wash buffer was removed by centrifugation for 1 min at 14 000 x g at RT. DNase I was added and after a 15-min incubation at RT, wash buffer was added, and samples were centrifuged for 1 min at 14 000 x g at RT. The wash step was repeated in total of three times. RNA was eluted by adding elution buffer to columns and centrifuging the samples for 1 min at 14 000 x g at RT. The eluates were incubated for 5 min at 65°C on a shaker followed by immediate cooling on ice. RNA concentration of each sample was measured using NanoDrop ND-1000 spectrophotometer (Thermo Fisher Scientific). The samples were stored at −80°C until use.

### 2.4. RNA sequencing

Of the 597 follow-up samples, 147 were not suitable for gene expression analysis. The gene expression profile of 450 samples was determined by bulk RNA sequencing at the University of Helsinki, FIMM Genomics unit.

3’ UTR-seq Library preparation was conducted as follows. The 3’ RNA-sequencing method was designed by Saavalainen-lab based on the Drop-seq protocol [42,43]. Briefly, 10 ng of RNA was mixed with Indexing Oligonucleotides (Integrated DNA Technologies, Coralville, IA, USA), 10 mM deoxynucleoside triphosphate (R0192, Thermo Scientific, Vilnius, Lithuania), RNAse inhibitor (EO0381, Thermo Scientific, Vilnius, Lithuania) and molecular grade water (Initial mix). After a 5-min incubation at ambient temperature, RNA was combined with reverse transcriptase mix containing 5× Maxima reverse transcriptase buffer, 200 U/µl Maxima reverse transcriptase (EP0742, Thermo Fisher Scientific), 40 U/µl Ribolock RNAse inhibitor (EO0381,Thermo Fisher Scientific), 50 µM Template Switch Oligo (231721686, Integrated DNA Technologies, Coralville, IA, USA) and 5 M betaine (J77507.VCR, Thermo Scientific). Samples were incubated in a T100 thermal cycler (Bio-Rad Laboratories Inc.) for 30 min at 22°C and 90 min at 52°C. The constructed complementary DNA was amplified by PCR in a volume of 30 μl using 10 μl of reverse transcriptase mix as template, 2× KAPA HiFi HotStart Readymix (07958935001, Kapa Biosystems, Wilmington, MA, USA), and 100 µM SMART PCR primer (234846524, Integrated DNA Technologies, Coralville, IA, USA). The samples were placed in a T100 thermocycler (Bio-Rad Laboratories Inc.) and run as follows: 95°C for 3 min; then 4 cycles of 98°C for 20 sec, 65°C for 45 sec, 72°C for 3 min; then 6 cycles of 98°C for 20 sec, 67°C for 20 sec, 72°C for 3 min; and with the final extension step of 5 min at 72°C and hold at 4°C. The PCR products were pooled together in sets containing different indexing oligos and purified with 0.6× Agencourt AMPure XP Beads (A63882, Beckman Coulter, Brea, CA, USA) according to the manufacturer’s instructions. The samples were eluted in 20 μl of molecular grade water. The 3’-end complementary DNA fragments for sequencing were prepared using the Illumina Nextera XT tagmentation reaction (15032350, Illumina, San Diego, CA, USA), with 1 ng of each PCR product pool serving as an input. The reaction was performed according to the manufacturer’s instructions, with the exception of the P5 SMART primer (231721688, Integrated DNA Technologies), which was used instead of the S5xx Nextera primer. Each set of samples that was pooled after the PCR reaction was tagmented with a different Nextera N7xx index (15052163, Illumina). Subsequently, the samples were PCR amplified as follows: 72°C for 3 min, 95°C for 30 sec, 12 cycles of 95°C for 10 sec, 55°C for 30 sec, and 72°C for 30 sec, with the final extension step of 5 min at 72°C and hold at 10°C. Samples were purified twice using 0.6× and 1.0× Agencourt AMPure Beads (A63882, Beckman Coulter) and eluted in 15 μl of molecular-grade water. The concentration of the libraries was measured using a Qubit 2 fluorometer (Invitrogen, Waltham, MA, USA) and the Quant-iT 1X dsDNA HS Assay Kit (Q33232, Thermo Fisher Scientific). The quality of the sequencing libraries was assessed using the TapeStation DNA High Sensitivity Assay (5067-5365, Agilent, Santa Clara, CA, USA). The libraries were sequenced on an Illumina NovaSeq 6000 instrument, S1 flow cells with a read 1 custom primer producing read 1 of 21 base pairs and read 2 of 106 base pairs.

### 2.5. Bioinformatic analyses

Read Alignment and Generation of Digital Expression Data was conducted as follows. The raw sequence data was inspected using FastQC (v0.11.9) [44] and MultiQC (v1.9) [45] to ensure the quality of sequencing results. Subsequently, the reads were filtered to remove low quality reads and reads shorter than 20 base pairs using Trimmomatic (v0.39) [46]. The reads passing the filter were then processed further using Drop-seq tools and the pipeline originally suggested in Drop-seq paper (v2.4.1) [42]. In brief, the raw, filtered read libraries were converted to sorted BAM files using Picard tools (https://broadinstitute.github.io/picard). This was followed by tagging reads with sample specific barcodes and unique molecular identifiers (UMIs). Tagged reads were then trimmed for 5’ adapters and 3’ poly-A tails. The alignment ready reads were converted from BAM formatted files to fastq files that were used as an input for STAR aligner (v2.7.6a) [47]. Alignments were done using data from Gencode Human Release 42 (GRCh38.p13) and comprehensive gene annotation files [48] with default STAR settings. Following the alignment, the uniquely aligned reads were sorted and merged with previous unaligned tagged BAM file to regain barcodes and UMI’s lost during the alignment step. Next, annotation tags are added to aligned and barcode tagged BAM files to complete the alignment process. Finally, Drop-seq tools are used to detect and correct systematic synthesis errors present in sample barcode sequences. Digital expression matrices were then created by counting the total number of unique UMI sequences (UMI sequences that differ only by a single base are merged) for each transcript.

DESeq2 (v1.38) was then used to identify differentially expressed genes [49]. Outlier samples were detected by running robust principal component analysis (rPCA) on DESeq2 normalized counts using PcaGrid function [50] implemented in the rrcov R package (v1.5-5) [51]. Sample was identified as an outlier using a cutoff value of 0.985 for computed orthogonal and score distances. The number of components included in the analysis was determined by scree test optimal coordinates and Kaiser criterion. This approach has been previously shown as an accurate approach for outlier sample detection in RNA-Seq data [52]. Differential expression analysis was then performed using Benjamini-Hochberg procedure to adjust for multiple testing. Additional annotations were added with biomaRt R package (v2.54.1).

We compared the global gene expression profiles of the follow-up gender-matched tertiles with Ingenuity Pathway Analysis (IPA) software v134816949 (Qiagen). Altered gene expression pathways were regarded significant at *p*<0.05. Out of these pathways, we included in our assessment the ones with a z-score over 2.0 indicating overexpression and the ones with a z-score below −2.0 indicating decreased gene expression. To enable the comparison of gene expression results in the further statistical analysis, we calculated the gene expression scores of each participant for the relevant pathways using RStudio v2024.04.1+748 software (RStudio, PBC, Boston, MA, USA), as previously described for the OXPHOS pathway score [53]. Similarly to the OXPHOS score, we defined a neutrophil degranulation (NeDegra) score based on genes included in this pathway in the IPA database and an identically calculated mtDNA score for the 13 protein-encoding genes of the mtDNA (S1-3 Tables).

### 2.6. Statistical analyses

We performed all statistical analyses with IBM SPSS Statistics, v29.0.1.0 software (IBM Corp., Amronk, NY, USA). We evaluated the distribution of different variables according to the Smirnov-Kolmogorov test. Continuous variables are described as their mean and standard deviation in the study population when normally distributed or median and 1^st^-3^rd^ interquartile range otherwise. For the comparison of normally distributed variables or variables that resulted in normal distribution after logarithmic transformation, we used the analysis of variance (ANOVA) method and Tukey’s post-hoc test for the transformed values. We compared non-normally distributed variables with the Kruskal-Wallis test and, when appropriate, the Mann-Whitney U-test. Gene expression scores were compared using the ANOVA method, as they were normally distributed. Categorical variables are shown as frequency and proportion, and were compared using Pearson’s Chi-Square test. When the assumptions of the Pearson’s Chi-Square test were violated (i.e., expected cell counts < 5), Fisher’s Exact Test was used. We tested the correlation between continuous variables with the Pearson product-moment correlation coefficient test. Results of all statistical tests were considered significant at *p*<0.05, unless stated otherwise.

Survival curves were generated with the Kaplan-Meier estimator. We assessed the differences of all-cause and cause-specific mortality among the blood mtDNA abundance tertiles based on the log-rank test. We used Cox proportional hazards regression analysis to calculate hazard ratios (HR) and 95% confidence intervals (CI) of mortality rates within population subgroups, while accounting for significant covariates. The proportional hazards assumption was evaluated using log-log survival plots. All figures were prepared using GraphPad Prism v10.3.1 (GraphPad Software, San Diego, CA, USA) software.

We defined the most relevant Cox regression adjustment model based on the characteristics of the sex-matched blood mtDNA tertiles of both the baseline and follow-up populations. All variables associated with the mtDNA tertiles at *p*<0.15 were considered as potential confounding factors. Variables that were significantly different without having an increasing or decreasing trend, were included in our assessment, if they showed a significant increasing or decreasing trend in either sex, assessed separately (S4-7 Tables). Next, we tested these confounding variables and removed the ones that were not significant when assessed in combination with others. In the baseline population, our final Cox regression adjustment model consisted of age, the presence of type 2 diabetes or CVD (including coronary heart disease and stroke [excluding subarachnoid hemorrhage]), left ventricular mass index and white blood cell count. For the follow-up cohort we included age, the presence of type 2 diabetes or pulmonary disease, and creatinine level into our final adjustment model.

## 3. Results

### 3.1. Description of study population

#### 3.1.1. Baseline

To examine associations between blood mtDNA abundance and clinical and epidemiological variables, we first divided our middle-aged baseline population into sex-matched tertiles based on the mtDNA abundance in their blood samples. This approach allowed fair comparison of the tertiles, as males had higher blood mtDNA abundance than females at baseline in our study population (S1A Fig). Tertile 1 represented the lowest, tertile 2 moderate and tertile 3 the highest amount of blood mtDNA, approximately 2-fold higher than tertile 1 (Table 1). The differences in blood mtDNA abundance across all tertiles were statistically significant, validating meaningful comparisons of these subpopulations.

**Table 1.** Comparison of continuous variables across blood mtDNA tertiles in the baseline and follow-up populations.

|  | Baseline |  |  |  | Follow-up |  |  |  |
| --- | --- | --- | --- | --- | --- | --- | --- | --- |
|  | Tertile 1 (n= 348) | Tertile 2 (n=345) | Tertile (n=330) | p-value | Tertile 1 (n=221) | Tertile 2 (n=183) | Tertile 3 (n=193) | p-value |
| Abundance of mtDNA | 0.58 (0.52-0.63) | 0.74 (0.68-0.79) | 0.96 (0.89-1.08) | <0.001<sup>1,3,5</sup> | 0.65 (0.59-0.69) | 0.77 (0.74-0.80) | 0.91 (0.86-0.96) | <0.001<sup>1,3,5</sup> |
| Age (years) | 52.5 (46.6-57.4) | 52.2 (47.6-56.6) | 50.0 (45.1-55.0) | <0.001<sup>1,3</sup> | 73.0 (69.0-78.0) | 72.0 (67.0-76.0) | 69.0 (66.0-74.0) | <0.001<sup>1,3</sup> |
| Height (cm) | 167.0 (160.5-175.0) | 167.0 (161.0-174.0) | 167.0 (161.0-175.0) | 0.515 | 165.0 (159.0-171.7) | 164.0 (157.8-172.0) | 164.5 (157.5-173.3) | 0.787 |
| Weight (kg) | 78.2 (67.7-88.5) | 73.7 (64.0-87.6) | 77.5 (67.2-87.3) | 0.020<sup>6</sup> | 78.9 (67.5-91.5) | 78.2 (69.2-87.8) | 76.4 (68.7-87.9) | 0.552 |
| Body mass index (kg/m<sup>2</sup>) | 27.4 (25.1-30.9) | 26.3 (24.0-29.8) | 27.1 (24.3-30.1) | 0.003<sup>5</sup> | 28.5 (25.7-32.3) | 28.2 (25.9-32.3) | 28.1 (25.3-31.3) | 0.427 |
| Waist circumference (cm) | 91.0 (82.0-101.0) | 89.0 (78.0-98.0) | 90.0 (81.0-98.0) | 0.031<sup>5</sup> | 97.0 (86.5-107.0) | 96.5 (88.5-106.5) | 94.5 (87.0-103.5) | 0.437 |
| Leptin (ng/ml) | 9.1 (5.2-13.7) | 7.7 (4.6-13.6) | 7.9 (5.1-14.2) | 0.254 | 15.5 (7.6-27.2) | 15.6 (7.7-33.3) | 14.7 (7.1-27.8) | 0.487 |
| Adiponectin (mg/l) | 15.0 (10.5-19.7) | 15.2 (11.1-19.7) | 15.0 (11.1-19.8) | 0.785 | 14.4 (10.5-19.4) | 13.0 (8.9-18.4) | 13.5 (10.0-18.6) | 0.158 |
| Resistin, only controls included (ng/ml) | 7.4 (5.9-9.3) | 7.1 (5.9-8.7) | 6.8 (5.7-8.8) | 0.322 | 10.9 (8.7-13.5) | 9.9 (8.3-13.4) | 9.6 (7.7-11.4) | 0.001<sup>1</sup> |
| Systolic blood pressure (mmHg) | 150 (134-166) | 147 (134-162) | 144 (129-158) | <0.001<sup>1</sup> | 138 (122-154) | 140 (123-154) | 137 (122-153) | 0.745 |
| Diastolic blood pressure (mmHg) | 91 (82-99) | 90 (82-97) | 88 (79-96) | 0.013<sup>2</sup> | 72 (64-80) | 73 (66-80) | 73 (67-80) | 0.525 |
| Left ventricular mass index (g/m<sup>2</sup>) | 130 (109-157) | 121 (101-150) | 122 (103-146) | 0.004<sup>2,5</sup> | 114 (97-134) | 115 (95-130) | 108 (93-129) | 0.125 |
| Fasting glucose (mmol/l) | 4.5 (4.1-4.9) | 4.4 (4.1-4.8) | 4.4 (4.1-4.7) | 0.137 | 6.0 (5.5-6.7) | 5.9 (5.4-6.8) | 5.9 (5.5-6.4) | 0.528 |
| Fasting insulin (mmol/l) | 11.7 (8.0-18.3) | 9.9 (7.0-14.7) | 10.3 (7.3-16.1) | 0.008<sup>5</sup> | 13.5 (9.1-20.6) | 13.5 (9.4-20.1) | 13.0 (9.2-17.8) | 0.440 |
| Quick insulin sensitivity check index | 0.59 (0.51-0.65) | 0.61 (0.54-0.68) | 0.60 (0.53-0.67) | 0.010<sup>5</sup> | 0.52 (0.47-0.58) | 0.52 (0.47-0.58) | 0.53 (0.48-0.58) | 0.324 |
| Homeostatic model assessment of insulin resistance | 2.27 (1.55-4.03) | 2.00 (1.34-3.11) | 2.03 (1.37-3.32) | 0.009<sup>5</sup> | 3.72 (2.36-5.93) | 3.77 (2.41-6.01) | 3.48 (2.28-5.18) | 0.324 |
| Non-alcoholic fatty liver disease fibrosis score | N/A | N/A | N/A | N/A | -0.1 (-0.8-0.8) | -0.2 (-0.9-0.5) | -0.4 (-1.2-0.5) | 0.007<sup>1</sup> |
| Smoking pack-years | 1.5 (0.0-18.0) | 1.0 (0.0-16.0) | 2.0 (0.0-16.0) | 0.551 | 0.0 (0.0-11.0) | 0.0 (0.0-15.0) | 0.0 (0.0-19.0) | 0.359 |
| Alcohol consumption (g/week) | 24.0 (2.0-84.0) | 24.0 (3.0-82.0) | 29.0 (3.0-84.0) | 0.544 | 12.0 (0.0-36.0) | 12.0 (0.0-48.0) | 24.0 (0.0-60.0) | 0.002<sup>1</sup> |
| Total cholesterol (mmol/l) | 5.7 (5.0-6.4) | 5.7 (5.0-6.3) | 5.7 (5.0-6.3) | 0.785 | 4.5 (3.9-5.3) | 4.6 (3.9-5.3) | 5.1 (4.2-5.5) | 0.009<sup>1</sup> |
| **High-density lipoprotein (mmol/l)** | 1.3 (1.1-1.6) | 1.3 (1.1-1.6) | 1.3 (1.1-1.5) | 0.239 | 1.4 (1.2-1.8) | 1.4 (1.2-1.7) | 1.4 (1.2-1.8) | 0.693 |
| **Low-density lipoprotein (mmol/l)** | 3.6 ± 0.9 | 3.5 ± 1.0 | 3.5 ± 0.9 | 0.855 | 2.9 ± 0.9 | 2.8 ± 0.9 | 3.0 ± 1.0 | 0.006<sup>1,4</sup> |
| **Triglycerides (mmol/l)** | 1.4 (1.0-1.9) | 1.3 (1.0-1.8) | 1.3 (1.0-1.9) | 0.422 | 1.1 (0.9-1.6) | 1.2 (0.9-1.6) | 1.2 (1.0-1.6) | 0.800 |
| **Creatinine (μmol/l)** | 81.0 (74.0-90.0) | 81.0 (72.0-89.0) | 80.0 (72.0-90.0) | 0.618 | 71.0 (61.0-86.0) | 69.0 (61.0-79.0) | 67.0 (59.0-77.0) | 0.021<sup>2</sup> |
| **Estimated glomerular filtration rate (ml/min/1.73m<sup>2</sup>)** | 78.8 (69.9-92.1) | 80.7 (71.0-91.7) | 80.5 (70.4-91.6) | 0.792 | 83.8 (70.0-90.5) | 85.9 (73.8-91.5) | 87.7 (78.9-93.5) | <0.001<sup>1</sup> |
| **High-sensitivity C-reactive protein (mg/l)** | 2.0 (0.8-5.2) | 1.4 (0.7-3.3) | 1.4 (0.7-3.0) | <0.001<sup>1,5</sup> | 1.4 (0.7-3.4) | 1.5 (0.6-3.3) | 1.2 (0.6-2.7) | 0.276 |
| **Alanine aminotransferase (U/l)** | 27.0 (20.0-38.0) | 26.0 (19.0-35.0) | 27.0 (20.0-36.0) | 0.147 | 23.0 (17.0-30.0) | 23.0 (18.0-31.0) | 24.0 (19.0-32.0) | 0.237 |
| **Hemoglobin (g/l)** | 142 (134-153) | 143 (132-151) | 143 (133-151) | 0.762 | 140 (132-149) | 140 (132-149) | 142 (133-150) | 0.505 |
| **White blood cell count (10<sup>9</sup>/l)** | 5.9 (5.0-7.2) | 5.6 (4.8-6.6) | 5.3 (4.5-6.2) | <0.001<sup>1,3,6</sup> | 5.8 (5.0-7.0) | 5.9 (4.8-7.0) | 5.4 (4.5-6.5) | 0.008<sup>1</sup> |
| **Platelet count (10<sup>3</sup>/l)** | 280 (237-323) | 277 (235-314) | 274 (236-317) | 0.756 | 220 (188-259) | 222 (192-257) | 226 (197-264) | 0.338 |
| **Telomere length** | 0.93 (0.81-1.08) | 0.91 (0.77-1.06) | 0.90 (0.79-1.06) | 0.295 | 0.54 (0.44-0.65) | 0.53 (0.44-0.62) | 0.51 (0.47-0.60) | 0.427 |

Data are presented as mean ± standard deviation (SD) or as median with 1<sup>st</sup>-3<sup>rd</sup> interquartile range. Interquartile ranges are shown in brackets. *P*-values were calculated using one-way ANOVA followed by Tukey's post hoc test or Kruskal-Wallis test followed by Mann-Whitney U-test. The tertiles of low, moderate and high blood mtDNA abundance of the follow-up cohort are, by definition, not a fraction of the respective baseline tertiles.
See Methods section for details.
1. *p*<0.01 between tertile 1 & 3.
2. *p*<0.05 between tertile 1 & 3.
3. *p*<0.01 between tertile 2 & 3.
4. *p*<0.05 between tertile 2 & 3.
5. *p*<0.01 between tertile 1 & 2.
6. *p*<0.05 between tertile 1 & 2.

We first compared the clinical characteristics typically associated with increased morbidity across the three tertiles. These adverse clinical profiles were significantly more profound in tertile 1, and included older age, higher systolic and diastolic blood pressure, higher high-sensitivity C-reactive protein levels, and higher white blood cell count (Table 1), as well as a higher prevalence of hypertension, type 2 diabetes and CVD (including coronary artery disease and cerebrovascular disease) (Table 2). All of these measures were the highest in tertile 1 and exhibited a progressive decline across the tertiles 2 and 3. Statistically significant differences in medication use were restricted to more frequent treatment of hypertension (*i.e.*, calcium channel blockers and angiotensin-converting enzyme blockers) in tertile 1 (Table 2), which may be the result of the previously described disease burden. However, some features associated with higher cardiovascular risk like body mass index, left ventricular mass index and fasting glucose, were significantly different across the tertiles (Table 1), but they did not show a consistent increasing or decreasing pattern from tertile 1 to 3. Next, we compared these clinical variables across blood mtDNA tertiles separately in males and females (S4-7 Tables). Left ventricular mass index was significantly different across the tertiles and showed a decreasing pattern from tertile 1 to 3 in males (S4 Table), and was therefore included in subsequent analyses as a confounding factor. Overall, our results suggest that lower blood mtDNA levels are linked to clinical profiles indicative of higher morbidity risk.

**Table 2.**
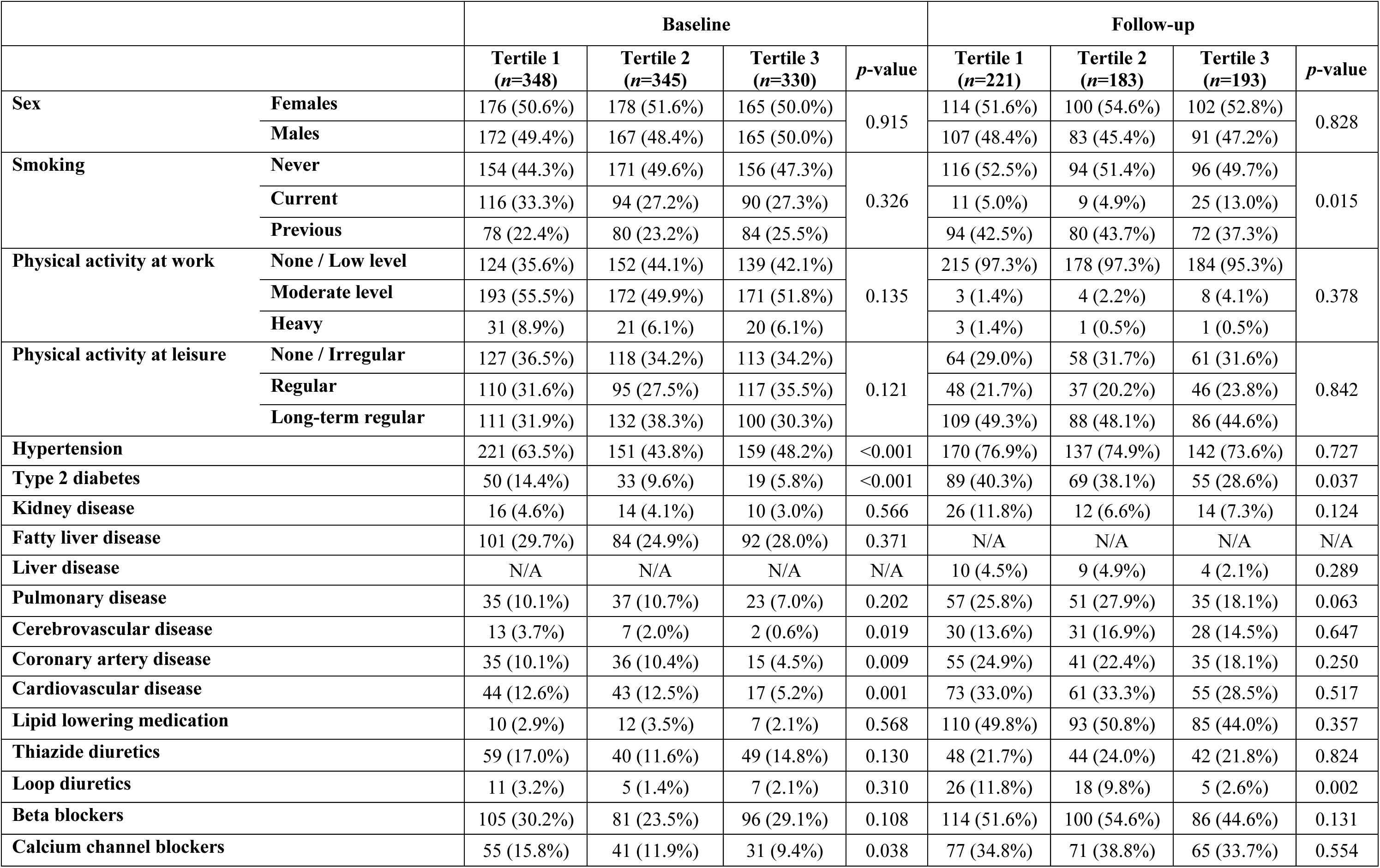
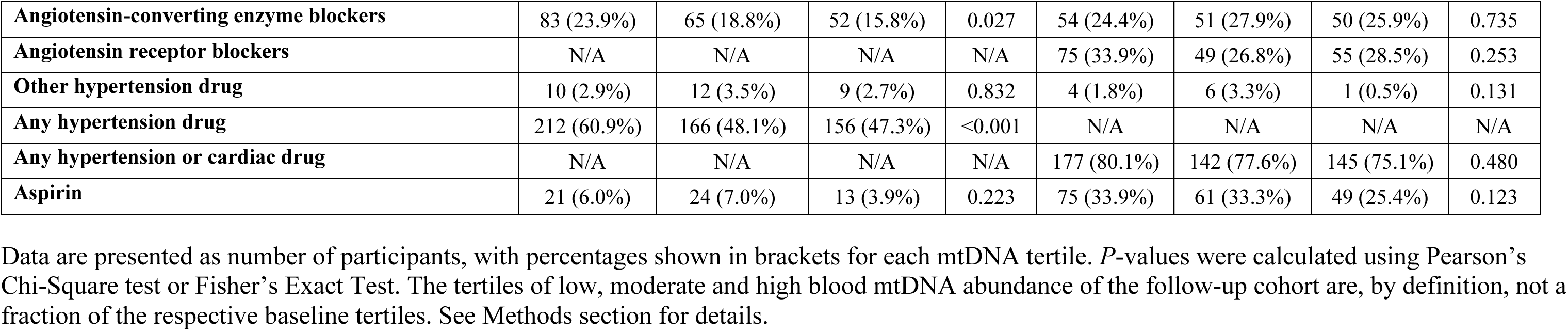
Comparison of categorical variables across blood mtDNA tertiles in the baseline and follow-up populations.

|  |  | Baseline |  |  |  | Follow-up |  |  |  |
| --- | --- | --- | --- | --- | --- | --- | --- | --- | --- |
|  |  | Tertile 1<br>(n=348) | Tertile 2<br>(n=345) | Tertile 3<br>(n=330) | p-value | Tertile 1<br>(n=221) | Tertile 2<br>(n=183) | Tertile 3<br>(n=193) | p-value |
| Sex | Females | 176 (50.6%) | 178 (51.6%) | 165 (50.0%) | 0.915 | 114 (51.6%) | 100 (54.6%) | 102 (52.8%) | 0.828 |
|  | Males | 172 (49.4%) | 167 (48.4%) | 165 (50.0%) |  | 107 (48.4%) | 83 (45.4%) | 91 (47.2%) |  |
| Smoking | Never | 154 (44.3%) | 171 (49.6%) | 156 (47.3%) | 0.326 | 116 (52.5%) | 94 (51.4%) | 96 (49.7%) | 0.015 |
|  | Current | 116 (33.3%) | 94 (27.2%) | 90 (27.3%) |  | 11 (5.0%) | 9 (4.9%) | 25 (13.0%) |  |
|  | Previous | 78 (22.4%) | 80 (23.2%) | 84 (25.5%) |  | 94 (42.5%) | 80 (43.7%) | 72 (37.3%) |  |
| Physical activity at work | None / Low level | 124 (35.6%) | 152 (44.1%) | 139 (42.1%) | 0.135 | 215 (97.3%) | 178 (97.3%) | 184 (95.3%) | 0.378 |
|  | Moderate level | 193 (55.5%) | 172 (49.9%) | 171 (51.8%) |  | 3 (1.4%) | 4 (2.2%) | 8 (4.1%) |  |
|  | Heavy | 31 (8.9%) | 21 (6.1%) | 20 (6.1%) |  | 3 (1.4%) | 1 (0.5%) | 1 (0.5%) |  |
| Physical activity at leisure | None / Irregular | 127 (36.5%) | 118 (34.2%) | 113 (34.2%) | 0.121 | 64 (29.0%) | 58 (31.7%) | 61 (31.6%) | 0.842 |
|  | Regular | 110 (31.6%) | 95 (27.5%) | 117 (35.5%) |  | 48 (21.7%) | 37 (20.2%) | 46 (23.8%) |  |
|  | Long-term regular | 111 (31.9%) | 132 (38.3%) | 100 (30.3%) |  | 109 (49.3%) | 88 (48.1%) | 86 (44.6%) |  |
| Hypertension |  | 221 (63.5%) | 151 (43.8%) | 159 (48.2%) | <0.001 | 170 (76.9%) | 137 (74.9%) | 142 (73.6%) | 0.727 |
| Type 2 diabetes |  | 50 (14.4%) | 33 (9.6%) | 19 (5.8%) | <0.001 | 89 (40.3%) | 69 (38.1%) | 55 (28.6%) | 0.037 |
| Kidney disease |  | 16 (4.6%) | 14 (4.1%) | 10 (3.0%) | 0.566 | 26 (11.8%) | 12 (6.6%) | 14 (7.3%) | 0.124 |
| Fatty liver disease |  | 101 (29.7%) | 84 (24.9%) | 92 (28.0%) | 0.371 | N/A | N/A | N/A | N/A |
| Liver disease |  | N/A | N/A | N/A | N/A | 10 (4.5%) | 9 (4.9%) | 4 (2.1%) | 0.289 |
| Pulmonary disease |  | 35 (10.1%) | 37 (10.7%) | 23 (7.0%) | 0.202 | 57 (25.8%) | 51 (27.9%) | 35 (18.1%) | 0.063 |
| Cerebrovascular disease |  | 13 (3.7%) | 7 (2.0%) | 2 (0.6%) | 0.019 | 30 (13.6%) | 31 (16.9%) | 28 (14.5%) | 0.647 |
| Coronary artery disease |  | 35 (10.1%) | 36 (10.4%) | 15 (4.5%) | 0.009 | 55 (24.9%) | 41 (22.4%) | 35 (18.1%) | 0.250 |
| Cardiovascular disease |  | 44 (12.6%) | 43 (12.5%) | 17 (5.2%) | 0.001 | 73 (33.0%) | 61 (33.3%) | 55 (28.5%) | 0.517 |
| Lipid lowering medication |  | 10 (2.9%) | 12 (3.5%) | 7 (2.1%) | 0.568 | 110 (49.8%) | 93 (50.8%) | 85 (44.0%) | 0.357 |
| Thiazide diuretics |  | 59 (17.0%) | 40 (11.6%) | 49 (14.8%) | 0.130 | 48 (21.7%) | 44 (24.0%) | 42 (21.8%) | 0.824 |
| Loop diuretics |  | 11 (3.2%) | 5 (1.4%) | 7 (2.1%) | 0.310 | 26 (11.8%) | 18 (9.8%) | 5 (2.6%) | 0.002 |
| Beta blockers |  | 105 (30.2%) | 81 (23.5%) | 96 (29.1%) | 0.108 | 114 (51.6%) | 100 (54.6%) | 86 (44.6%) | 0.131 |
| Calcium channel blockers |  | 55 (15.8%) | 41 (11.9%) | 31 (9.4%) | 0.038 | 77 (34.8%) | 71 (38.8%) | 65 (33.7%) | 0.554 |

|  |  |  |  |  |  |  |  |  |
| --- | --- | --- | --- | --- | --- | --- | --- | --- |
| **Angiotensin-converting enzyme blockers** | 83 (23.9%) | 65 (18.8%) | 52 (15.8%) | 0.027 | 54 (24.4%) | 51 (27.9%) | 50 (25.9%) | 0.735 |
| **Angiotensin receptor blockers** | N/A | N/A | N/A | N/A | 75 (33.9%) | 49 (26.8%) | 55 (28.5%) | 0.253 |
| **Other hypertension drug** | 10 (2.9%) | 12 (3.5%) | 9 (2.7%) | 0.832 | 4 (1.8%) | 6 (3.3%) | 1 (0.5%) | 0.131 |
| **Any hypertension drug** | 212 (60.9%) | 166 (48.1%) | 156 (47.3%) | <0.001 | N/A | N/A | N/A | N/A |
| **Any hypertension or cardiac drug** | N/A | N/A | N/A | N/A | 177 (80.1%) | 142 (77.6%) | 145 (75.1%) | 0.480 |
| **Aspirin** | 21 (6.0%) | 24 (7.0%) | 13 (3.9%) | 0.223 | 75 (33.9%) | 61 (33.3%) | 49 (25.4%) | 0.123 |

Data are presented as number of participants, with percentages shown in brackets for each mtDNA tertile. *P*-values were calculated using Pearson's Chi-Square test or Fisher's Exact Test. The tertiles of low, moderate and high blood mtDNA abundance of the follow-up cohort are, by definition, not a fraction of the respective baseline tertiles. See Methods section for details.

#### 3.1.2. Follow-up

To assess whether the findings of the baseline population extend to the more elderly follow-up cohort, we stratified the available 597 returning subjects into tertiles based on their blood mtDNA levels. Although the blood mtDNA levels of males and females were comparable in the follow-up population (S1B Fig), we created sex-matched tertiles to maintain consistency with the baseline assessment. These follow-up tertiles are, by definition, not a fraction of the respective baseline tertiles, *i.e*., a subject’s tertile classification was determined based on the blood mtDNA abundance measured in their 2010 sample, regardless of which tertile the same subject was included in at baseline. Similar to baseline findings, tertile 1 exhibited the lowest and tertile 3 the highest blood mtDNA levels. However, in this elderly population, the amount of blood mtDNA in tertile 3 was approximately 1.5-fold higher than in tertile 1. Despite the narrower blood mtDNA range in comparison to baseline, significant differences were still observed across all tertiles in the follow-up population (Table 1).

During follow-up, lower blood mtDNA levels were again associated with clinical characteristics indicative of elevated morbidity risk, consistent with baseline observations. We found that older age, higher white blood cell count (Table 1) and the higher prevalence of type 2 diabetes (Table 2) were again characteristics of tertile 1. Additionally, tertile 1 had higher creatinine and accordingly lower estimated glomerular filtration rate, higher resistin levels and higher non-alcoholic fatty liver disease score (Table 1). Pulmonary disease tended to be more prevalent in tertile 1 (Table 2) and showed a decreasing pattern from tertile 1 to 3 in males (Sex-specific information is provided in S4-7 Tables). Therefore, it was included in subsequent analyses as a confounding factor. Unhealthier smoking habits (Table 2) were also characteristic of tertile 1, but interestingly, the proportion of current smokers was higher in tertile 3 (Table 2). Regarding medication, only loop diuretics were more frequent in tertile 1 than other tertiles (Table 2). As with the baseline analysis, generally these results reflected that the disease burden exhibited a decreasing pattern across the tertiles, with tertile 1 having the highest morbidity risk.

In contrast to previous results, we also identified features of negative clinical outcomes that were more substantial in tertile 3. Interestingly, these included trends of higher alcohol consumption, higher total cholesterol and higher LDL cholesterol (Table 1). We asked whether the differences in cholesterol levels could be explained by medication use. However, the proportion of individuals on lipid-lowering therapy was comparable across the tertiles. Nonetheless, even with small differences in CVD prevalence and medication use, secondary prevention was likely more efficient in the subjects of tertile 1, which can explain the findings regarding cholesterol levels.

In summary, some clinical characteristics associated with the baseline blood mtDNA tertiles, particularly in tertile 1, were attenuated with aging by the time of the follow-up examination, while new differences emerged over the 20 years since the baseline visit. These findings suggest that the relationship between blood mtDNA levels and clinical features can shift with age, thereby supporting the rationale for assessing blood mtDNA separately at different age groups in our study design.

### 3.2. All-cause mortality

#### 3.2.1. Baseline

After describing the clinical features of the blood mtDNA-based tertiles, we next examined the association of blood mtDNA levels with all-cause mortality. We registered a total of 189, 151 and 102 death events up to the end of 2022 in the sex-matched low, moderate and high blood mtDNA tertiles, respectively, with the proportion of death events decreasing towards higher blood mtDNA levels (Table 3). Pairwise comparisons across the three tertiles using the log-rank test revealed significantly different mortality rates. Subjects in the lowest blood mtDNA tertile (tertile 1) exhibited the highest mortality rate, followed by those with moderate mtDNA levels (tertile 2), while subjects in the highest mtDNA tertile (tertile 3) had the lowest mortality. These differences are illustrated by the Kaplan–Meier survival curves shown in Fig 1A. In the Cox proportional hazards analysis, we defined tertile 3 with high blood mtDNA abundance as the reference and applied the adjustment model described in the Statistical analysis section. The results showed a significant overall difference across tertiles, with participants in the lowest blood mtDNA group (tertile 1) having a 57% higher risk of all-cause mortality, and those in the moderate blood mtDNA tertile (tertile 2) having a 30% higher risk compared to those in the highest blood mtDNA tertile (tertile 3) (Fig 1A, S8 Table). The addition of hypertension to the Cox regression adjustment model, as a well-known factor influencing mortality [54] or the inflammatory marker high-sensitivity C-reactive protein, did not change the results substantially (S8 Table). Overall, our findings demonstrate that lower blood mtDNA levels are associated with a higher risk of all-cause mortality.

**Fig 1.**
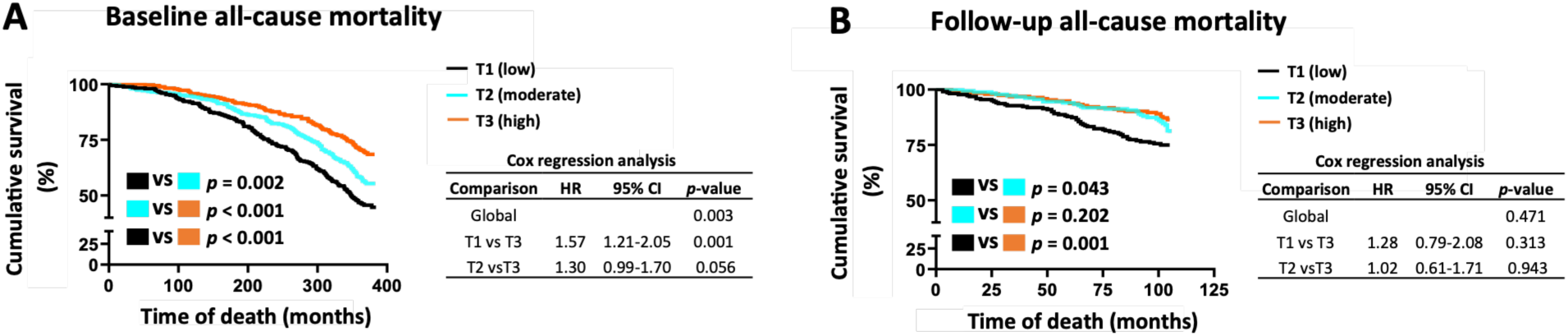
Lower blood mtDNA abundance is associated with higher all-cause mortality. (A) Baseline (*n*=1023) and (B) follow-up (*n*=597) Kaplan-Meier survival curves stratified by blood mtDNA tertiles: low (T1), moderate (T2) and high (T3). Pairwise differences between survival curves were assessed using the log-rank test. Results from the Cox proportional hazards regression analyses are shown to the right of each panel. At baseline (A), the adjustment model consisted of age, the presence of type 2 diabetes or cardiovascular diseases, left ventricular mass index and white blood cell count, while at follow-up (B), age, the presence of type 2 diabetes or pulmonary disease, and creatinine level were included. In (A), *n*=348 in T1, *n*=345 in T2 and *n*=330 in T3. In (B), *n*=221 in T1, *n*=183 in T2 and *n*=193 in T3. Detailed results from the Cox regression analyses are provided in S8 Table. HR, hazard ratio; CI, confidence interval.

**Table 3.** All-cause mortality across blood mtDNA tertiles.

|  | Baseline (n=1023) |  |  | Follow-up (n=597) |  |  |
| --- | --- | --- | --- | --- | --- | --- |
|  | Tertile 1 | Tertile 2 | Tertile 3 | Tertile 1 | Tertile 2 | Tertile 3 |
| Alive | 159 (45.7%) | 194 (56.2%) | 228 (69.1%) | 165 (74.7%) | 150 (82.0%) | 167 (86.5%) |
| Deceased | 189 (54.3%) | 151 (43.8%) | 102 (30.9%) | 56 (25.3%) | 33 (18.0%) | 26 (13.5%) |
| Total | 348 (100%) | 345 (100%) | 330 (100%) | 221 (100%) | 183 (100%) | 193 (100%) |

Data are presented as number of participants, with percentages shown in brackets for each mtDNA tertile. The tertiles of low, moderate and high blood mtDNA abundance of the follow-up cohort are, by definition, not a fraction of the respective baseline tertiles. See Methods section for details.

#### 3.2.2. Follow-up

Similar to baseline, we evaluated the association between blood mtDNA levels and all-cause mortality in the follow-up population. Among the subjects who returned for a second visit, the low, moderate, and high blood mtDNA tertiles recorded 56, 33, and 26 death events, respectively, by the end of 2022 levels (Table 3). This pattern reflected a consistent decreasing trend in mortality with increasing blood mtDNA levels. Pairwise comparisons with the log-rank test revealed that subjects in the lowest blood mtDNA tertile (tertile 1) had a significantly increased mortality rate, while subjects in the moderate and high blood mtDNA tertiles (tertiles 2-3) exhibited more comparable mortality outcomes (Fig 1B). The Cox proportional hazards analysis, conducted using the model described in the Statistical analysis section, showed no significant overall or pairwise differences in mortality risk among tertiles (Fig 1B, S8 Table). Including white blood cell count in the Cox regression adjustment model to align with the baseline analysis led to a similar nonsignificant result (S8 Table). In summary, we found that lower blood mtDNA levels are associated with a higher risk of mortality in the follow-up population. However, this association was attenuated to nonsignificant levels after adjusting for confounding factors identified in the Cox regression analysis. In conclusion, we found that in the more elderly follow-up population, the predictive power of blood mtDNA abundance for mortality was weaker compared to the younger baseline population.

### 3.3. Cause-specific mortality and cardiovascular morbidity

To describe blood mtDNA abundance as a potential predictive clinical biomarker for CVD or other leading causes of death in Western populations, we next examined the association of blood mtDNA abundance with cause-specific mortality up to the end of 2021. For this, we created a variable reflecting the cause of death events with three categories: “cardiovascular”, “cancer” and “other” causes, along with a fourth category, “alive”. We analyzed the distribution and time of death events across the blood mtDNA tertiles of the 1990s baseline and 2010s follow-up cohorts. For a comprehensive cardiovascular assessment, we conducted a separate analysis on the first recorded cardiovascular events, both fatal and non-fatal, experienced by the baseline study population up to the end of 2014.

#### 3.3.1. Cause-specific mortality differences at baseline

All death event categories followed a decreasing pattern towards higher blood mtDNA levels, and each cause of death accounted for approximately the same proportion to the total number of deaths. In the low, moderate, and high blood mtDNA tertiles, CVD accounted for 60, 45, and 31 deaths, cancer for 57, 35, and 34 deaths, while other causes for 63, 54, and 28 deaths, respectively (Fig 2A).

**Fig 2.**
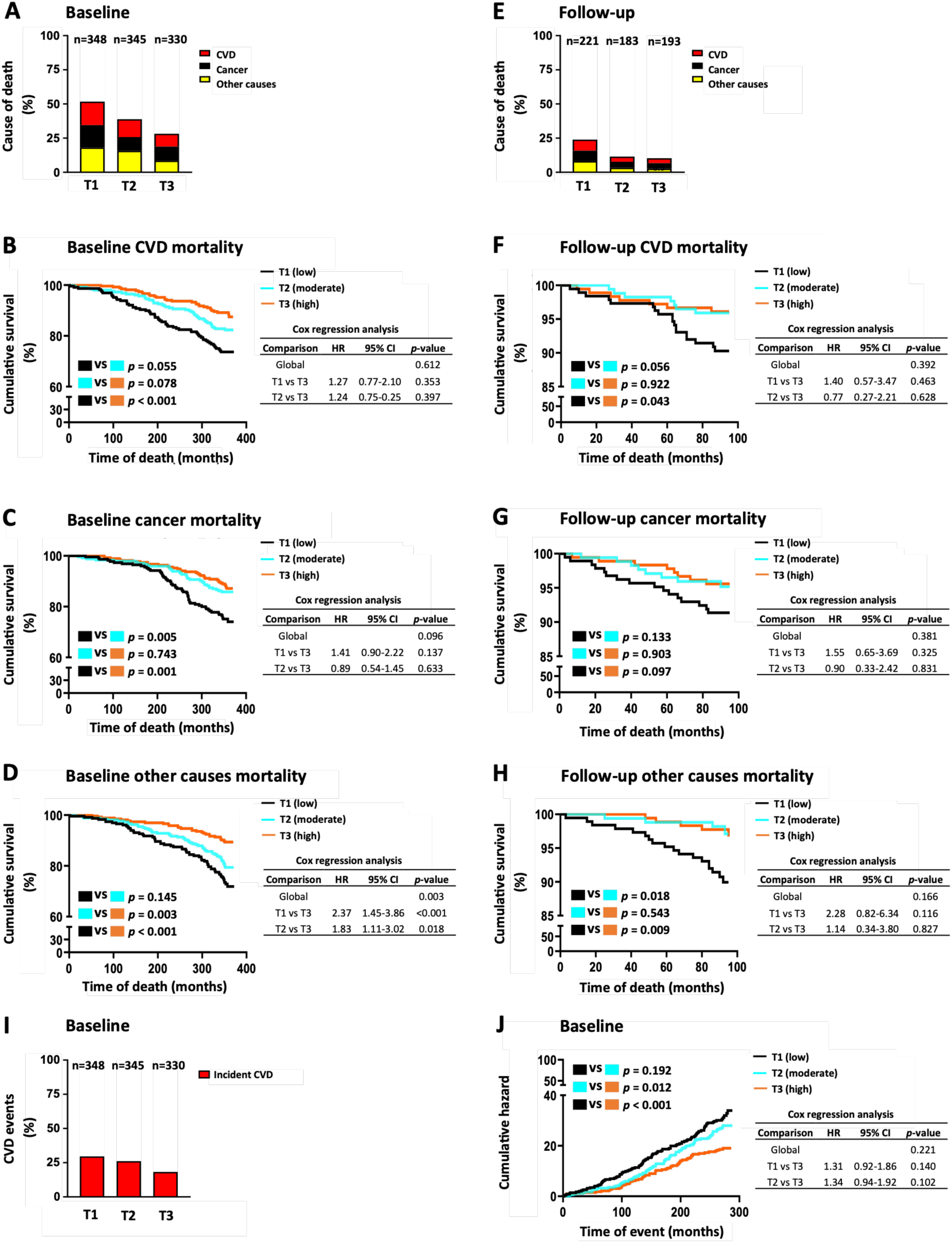
Lower blood mtDNA abundance is associated with higher cause-specific mortality and CVD morbidity. (A) Proportional distribution of death causes at baseline. (B-D) Kaplan-Meier survival curves for CVD (B), cancer (C) and other causes (D) at baseline. (E) Proportional distribution of death causes at follow-up. (F-H) Kaplan-Meier survival curves for CVD (F), cancer (G) and other causes (H) at follow-up. (I) Proportional distribution of incident CVD events at baseline. (J) Hazard curves for CVD events at baseline, depicting the instantaneous rate of events over time. Survival and hazard curves were stratified by blood mtDNA tertiles: low (T1), moderate (T2) and high (T3). Pairwise differences were assessed using the log-rank test. Results from the Cox proportional hazard regression analyses are shown to the right of each panel. At baseline (A-D and I-J), the adjustment model consisted of age, the presence of type 2 diabetes or cardiovascular diseases, left ventricular mass index and white blood cell count, while at follow-up (E-H) age, the presence of type 2 diabetes or pulmonary disease, and creatinine level were included. In (A-D and I-J), *n*=348 in T1, *n*=345 in T2 and *n*=330 in T3. In (E-H), *n*=221 in T1, *n*=183 in T2 and *n*=193 in T3. Detailed results from the Cox regression analyses are provided in S9 Table. CVD, cardiovascular diseases; HR, hazard ratio; CI, confidence interval.

The pairwise comparison of survival curves in the blood mtDNA tertiles using the log-rank test revealed that the low and high blood mtDNA tertiles were significantly different from each other in all death categories, with high blood mtDNA levels appearing as a protective factor (Fig 2B-D). In the case of “cancer”, the moderate blood mtDNA group (tertile 2) exhibited a mortality rate comparable to that of the high blood mtDNA group (tertile 3) within our study population. In the case of “cardiovascular” and “other” causes, subjects with moderate blood mtDNA levels (tertile 2) exhibited risk levels in-between low and high blood mtDNA groups (tertile 1 and 3). Cox proportional hazards analysis revealed a significant overall difference across tertiles for “other” causes of death (Fig 2D, S9 Table), but not for “cardiovascular” or “cancer” (Fig 2B-C, S9 Table). For the “other” causes category, subjects with low blood mtDNA abundance (tertile 1) had more than 2-fold higher mortality risk, while those with moderate blood mtDNA abundance (tertile 2) had 83% higher risk compared to subjects with high blood mtDNA levels (tertile 3) (Fig 2D, S9 Table). In summary, our results indicated that lower blood mtDNA levels are associated with a higher risk of mortality across all the studied death categories. Notably, the association with death classified under “other” causes remained significant even after adjustment for the identified confounding factors.

#### 3.3.2. Cause-specific mortality differences at follow-up

We conducted a similar analysis across the elderly follow-up population to assess whether the same patterns in the causes of death could be revealed as during baseline. Similarly to the baseline results, each cause of death contributed approximately the same proportion of events to the overall mortality (Fig 2E). Although statistical power was limited due to the low number of death events in each category, the pairwise comparison of mtDNA tertiles showed similar patterns as baseline findings, with the low mtDNA tertile (tertile 1) consistently showing the highest number of deaths in each category (Fig 2E-H). Cox proportional hazards analysis demonstrated no significant overall or pairwise differences in mortality risk among tertiles (Fig 2F-H, S9 Table). Overall, these observed patterns suggest that lower blood mtDNA levels may be linked to increased cause-specific mortality risk across all examined categories also in the the follow-up population (Fig 2F-H). However, the association did not reach statistical significance after adjustment for potential confounders, which is not surprising given the analysis’ limited power.

#### 3.3.3. Cardiovascular event differences at baseline

To investigate the relationship between blood mtDNA and CVD in our population, we conducted a separate analysis in the statistically powered baseline cohort on all cardiovascular events, including the ones leading to death and hospitalization of a subject. CVD event data were available up to the end of 2014 (for detailed description of CVD events see Methods section), therefore, this analysis was not performed in the follow-up cohort. The proportion of subjects experiencing these cardiovascular events presented a decreasing trend in the blood mtDNA tertiles, as shown in Fig 2I. Pairwise comparisons of the tertiles using the log-rank test revealed significant differences in the risk of developing CVD events, with high blood mtDNA levels again exhibiting a protective effect (Fig 2J). No significant overall or pairwise differences in the risk of CVD events were observed across tertiles in the Cox proportional hazards analysis (Fig 2J, S9 Table). In summary, lower blood mtDNA levels are associated with the higher risk of developing CVD events. Nonetheless, this association was largely attenuated after adjusting for the identified confounding factors.

### 3.4. Gene expression profiles in the follow-up population

To gain insight into the molecular mechanisms that may influence blood mtDNA content and contribute to the observed mortality differences across the mtDNA tertiles, we performed RNA sequencing on 450 available buffy coat samples from the follow-up population. Our aim was to identify gene expression patterns and pathways associated with differing blood mtDNA levels between tertiles.

We conducted pairwise comparisons of the three sex-matched blood mtDNA tertiles with the IPA using RNA sequencing data adjusted for batch effects (S10-12 Tables). Interestingly, pathways related to immune cell activity showed the most prominent differences across all tertile comparisons, with neutrophil degranulation emerging as the most upregulated pathway at lower blood mtDNA level tertiles 1 and 2 (Fig 3A). Neutrophil degranulation, recognized as a hallmark of innate immune activation, likely contributes not only to acute inflammation but also plays a key role in sustaining the persistent, low-grade inflammation that characterizes cardiometabolic diseases [55]. To compare neutrophil degranulation across blood mtDNA tertiles, we calculated the neutrophil degranulation (NeDegra) gene expression score based on the IPA pathway gene list (S2 Table). The NeDegra score differed significantly across all tertiles (Fig 3B) and was significantly associated with an increased mortality risk (Fig 3C), with higher scores linked to greater risk. This association remained significant in the Cox regression analysis after adjusting for the confounding factors identified for the blood mtDNA tertiles (S13 Table). These findings suggest a potential link between blood mtDNA and inflammation, which we explored by analysing the correlation between mtDNA abundance and the NeDegra score. Indeed, we observed a moderate significant negative correlation (Fig. 3D). Together, these results support the concept that blood mtDNA content and neutrophil activation are interlinked, and their dysregulation may jointly contribute to disease progression.

**Fig 3.**
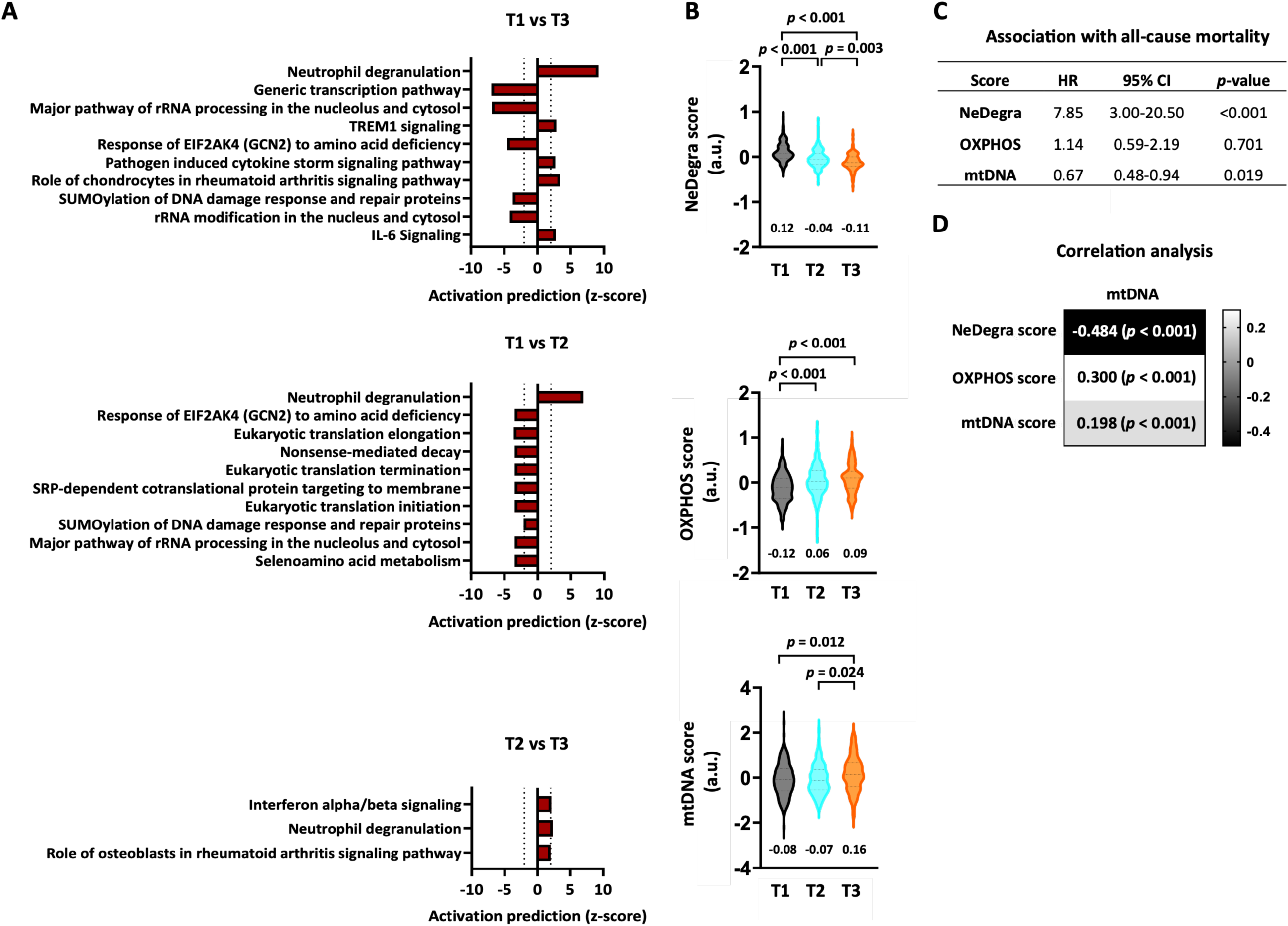
Transcriptomic analyses link blood mitochondrial DNA (mtDNA) abundance to neutrophil degranulation in the follow-up population. (A) The top ten most significantly altered pathways using differentially expressed global genes according to the Ingenuity Pathway Analysis (IPA) across the blood mtDNA tertiles with low (T1), moderate (T2) and high (T3) blood mtDNA abundance. Z-scores provided by IPA represent activation directionality of the pathways, with positive observed z-scores predicting “activated” and negative z-scores “inhibited” pathways. Analysis was carried out using genes at *p* < 0.05 and |log₂ fold change| > 0.2. Pathways with z-scores 2 or < −2 were considered statistically significant and only these pathways are shown. Pathways are ranked according to statistical significance. Notably, only three pathways were significantly altered between T2 and T3. (B) Transcriptomics-based analysis of neutrophil degranulation (NeDegra) score, oxidative phosphorylation (OXPHOS) score and mtDNA score across the blood mtDNA tertiles. Mean value of each tertile is shown below the violin plots. Gene lists used to calculate each gene expression score are provided in S1-3 Tables. (C) Association of gene expression scores with all-cause mortality, estimated using the Cox proportional hazards regression. (D) Heatmap of Pearson correlation coefficients between mtDNA abundance and gene expression scores. *P*-values are shown in brackets. In (A-D), *n*=173 in T1, *n*=132 in T2 and *n*=135 in T3. HR, hazard ratio; CI, confidence interval.

Lastly, to explore whether blood mtDNA levels are linked to blood cells’ OXPHOS capacity at the transcriptional level, we calculated two gene expression scores: an OXPHOS score [53], comprising nuclear-encoded genes of OXPHOS subunits, and an mtDNA score, containing the 13 protein-coding genes encoded by the mtDNA (S1 and S3 Tables). Both scores differed across the follow-up blood mtDNA tertiles, with tertile 1 showing the lowest values for both scores (Fig 3B). The OXPHOS score was not significantly associated with mortality in any model, whereas the mtDNA score showed a significant association (Fig 3C), which did not persist after adjustment in the Cox regression model (S13 Table). Furthermore, only weak positive correlations were observed between blood mtDNA abundance and either of these two gene expression scores (Fig 3D), suggesting that blood mtDNA content only weakly reflects blood cell OXPHOS or mtDNA related transcriptional activity.

In summary, blood mtDNA content showed the strongest correlation with the NeDegra score, suggesting that variations in blood mtDNA levels are closely linked to inflammatory activity. Moreover, low blood mtDNA abundance likely reflects underlying immune-metabolic dysregulation, with neutrophil activation emerging as a relevant mediator of mortality risk in the elderly follow-up population.

## 4. Discussion

Blood mtDNA levels are increasingly used as a marker of mitochondrial health [5]. Previous studies have associated blood mtDNA with major chronic diseases, but these studies typically have follow-up periods shorter than 20 years [35], and the disease specificity and pathomechanisms behind these associations remain unclear. Our study addresses these gaps by demonstrating that low blood mtDNA levels predict higher morbidity and mortality over a 30-year follow-up, with associations not limited to specific causes of death. Importantly, blood gene expression analysis revealed that low blood mtDNA abundance likely reflects underlying immune-metabolic dysregulation, which may contribute to the increased nonspecific mortality risk observed. Overall, our findings support blood mtDNA as a valuable biomarker for early risk assessment and primary prevention of various diseases.

Blood mtDNA levels are known to be influenced by a range of physiological and pathological factors, including age, sex, inflammatory status, metabolic health, and underlying disease conditions [22]. While previous studies have reported higher blood mtDNA in females [7,22,26,32,33], our middle-aged baseline cohort showed significantly higher mtDNA levels in males. This unexpected finding may reflect lifestyle differences in Finland during the 1980s–1990s or sampling variability. Thus, to control for potential sex-related bias, we used sex-matched tertiles in our analyses. As reported previously [7,17,22,23,31,32,34], older age, particularly above 50 years, was associated with lower blood mtDNA levels, and we observed a clear age-related decline across mtDNA tertiles. Consistent with earlier research [7,9,17–19,22–24,26], lower blood mtDNA levels were linked to adverse clinical profiles, including cardiovascular diseases and related risk factors such as obesity, type 2 diabetes, dyslipidemia and hypertension. Lower blood mtDNA was also associated with elevated inflammatory markers, such as higher white blood cell counts and high-sensitivity C-reactive protein levels, particularly during the baseline phase, similarly as previously shown [7–11,21]. Interestingly, in the elderly follow-up cohort, the highest blood mtDNA tertile included a greater proportion of current smokers and individuals with higher cholesterol levels, findings likely explained by more effective secondary prevention in other tertiles with higher disease burden. Alcohol consumption was also highest in tertile 3, especially among males, which may reflect greater overall well-being in this group, as frailty can limit alcohol use in older age. Altogether, these partly contrasting findings can likely be explained by the clinical aspects and lifestyle of the subjects in different tertiles and they did not appear to influence subsequent mortality analysis. Some differences in blood mtDNA levels observed at baseline were less pronounced in the older follow-up cohort. This attenuation may be explained by age-related changes in mtDNA dynamics, lifestyle differences between the age groups, and the smaller sample size in the follow-up phase. Overall, lower blood mtDNA levels remained consistently associated with clinical profiles linked to higher morbidity risk.

Blood mtDNA levels have previously been shown to predict all-cause mortality in several prospective epidemiological studies of European [21,31,34,36], American [32] and Asian [11,33] origin in different types of populations, with follow-up times of up to 17 years. Although the methodology of these types of studies is, by definition, not primarily suitable for establishing cause-specific relationships between blood mtDNA levels and mortality, they highlight the sensitivity of blood mtDNA in predicting mortality across different populations and circumstances. Our findings align with these earlier reports and extend them by demonstrating similar associations over a 30-year follow-up. This relationship appeared to be stronger in the middle-aged subjects of the baseline cohort, which can be explained by the larger number of cases and more heterogenous study population. In contrast, we found that in the more elderly follow-up cohort the differences in the ability of blood mtDNA abundance to predict mortality was weaker. Regarding this, it is important to recognize that many individuals with low blood mtDNA levels had died during follow-up, making the surviving elderly cohort more homogeneous and reducing the predictive value of blood mtDNA at older age. Nevertheless, this predictive value remains clinically relevant, as it offers a timeframe suitable for primary prevention. In summary, our results suggest that blood mtDNA levels in middle-aged individuals can predict upcoming all-cause mortality over a span of 30 years and can be used as a repeatedly quantified biomarker of upcoming morbidity.

Previous studies have reported associations between lower blood mtDNA levels and increased CVD risk [18–20,24,28]. In line with this, our baseline study population showed a higher prevalence of CVD among individuals with lower blood mtDNA levels. However, this association was no longer significant in the elderly follow-up cohort, possibly due to the more homogenous nature of the follow-up population, as discussed earlier. Notably, comprehensive genome sequencing and Mendelian randomization-based studies by Liu et al. [20] and Qin et al. [28] found no evidence for a causal relationship between low blood mtDNA levels and the prevalence of CVD. Regarding mortality, our primary assessment found that lower baseline blood mtDNA levels were associated with a higher CVD mortality and both fatal and non-fatal CVD events. However, these associations were attenuated to non-significant levels after adjusting for identified confounding factors. Overall, our results, together with evidence from previous studies, suggest that low blood mtDNA levels are linked to an elevated risk of CVD mortality over three decades and CVD events over two decades, but the relationship is unlikely to be causal.

The relationship between blood mtDNA and non-cardiovascular mortality or morbidity has been explored with varying focus and outcomes in the literature. Previous literature has often focused on single causes of death [21,30–34,36], representing a more selective approach compared to our comprehensive analysis, and producing inconsistent results. While some smaller cohort studies reported associations between low blood mtDNA levels and increased cancer-mortality [21,34], a large Mendelian randomization study using the FinnGen cohort [37] found no cause-specific relationship between blood mtDNA abundance and the risk of developing cancer. In our baseline cohort analysis, cancer-specific mortality was highest among individuals with lower blood mtDNA levels, but it lost significance after adjusting for confounders. Conversely, mortality from “other” causes was the most pronounced and showed a consistent association with low blood mtDNA, even after adjustment. Overall, these findings indicate that while blood mtDNA abundance may predict upcoming mortality, it lacks specificity for any particular cause of death. Instead, it may reflect various underlying pathological conditions in the body. This nonspecific predictive ability supports the utility of blood mtDNA as a general biomarker of systemic health, complementing its role in CVD risk prediction, as highlighted in a recent comprehensive meta-analysis [35].

Low blood mtDNA levels have also been consistently linked to systemic inflammation and elevated white blood cell counts [7–11,21]. Our gene expression analysis reinforces this interplay, particularly highlighting inflammatory pathways involving neutrophils, the most abundant white blood cell type. Although neutrophil degranulation is classically associated with acute inflammation, it also contributes to the chronic low-grade inflammatory state characteristic of cardiometabolic diseases [55]. In such conditions, sustained neutrophil activation may reduce blood mtDNA abundance through two complementary mechanisms. First, a shift in leukocyte composition, *i.e.,* an increased neutrophil-to-lymphocyte ratio, can lower total blood mtDNA amount, as neutrophils contain up to 3-fold less mtDNA than lymphocytes [56,57]. This is consistent with our mortality findings, as neutrophil-to-lymphocyte ratio has been linked to increased mortality in several studies [58–60]. Second, chronic exposure to neutrophil-derived mediators, such as reactive oxygen species and proteolytic enzymes, can damage mitochondrial structures, impair mtDNA replication, and disrupt mitochondrial biogenesis pathways by downregulating a key regulator of mitochondrial metabolism, peroxisome proliferator-activated receptor-γ coactivator 1α [61,62], ultimately leading to a progressive reduction in mtDNA content in circulating blood cells. Importantly, low blood mtDNA amount may not only result from inflammation but may also promote it. Rather than directly inducing inflammation, mitochondrial dysfunction associated with reduced blood mtDNA amount can promote neutrophil activation and chronic inflammation through increased reactive oxygen species production and disrupting anabolic and metabolic signalling pathways [63,64]. Interestingly, in our follow-up population, blood mtDNA content did not correlate well with the transcriptional activity of OXPHOS or mtDNA genes, which are central to ATP production. Instead, this association was stronger with the NeDegra score, which, unlike blood mtDNA abundance, also independently predicted mortality after adjustment. This observation suggests that variation in blood mtDNA abundance may be more tightly linked to inflammatory activity and mitochondrial functions beyond energy production in chronic cardiometabolic conditions. Together, these findings support the presence of a bidirectional interplay between mitochondria and innate immune activation, which may explain the observed differences in blood mtDNA content across tertiles and the nonspecific association between low blood mtDNA abundance and long-term morbidity and mortality. The persistent predictive value of the NeDegra score and its correlation with blood mtDNA levels suggest that neutrophil-driven inflammation may be a key mediator of this link.

### 4.1. Strengths

The key strengths of our study include a well-characterized population, a long 30-year follow-up period, and a longitudinal study design. Although the baseline cohort was recruited based on hypertension status, a high prevalence of hypertension in Finland in 1992 (>40% in this age group) [65] supports the generalizability of our findings to the middle-aged Finnish and broader Western populations. To the best of our knowledge, this is the first epidemiological study to demonstrate that the predictive value of blood mtDNA abundance, previously shown in studies with ≤17-year follow-up, extends up to 30 years in a general population. This extended follow-up time is particularly valuable for enabling early primary prevention, when the prevalence of chronic diseases is still low. Furthermore, the return of most living participants for a second visit after 20 years allowed us to assess the same associations at older age with reduced variability from external factors. In addition, our comprehensive cause-of-death assessment across all recorded deaths confirmed that blood mtDNA abundance lacks specificity for any single disease. An additional strength is the integration of blood gene expression analysis with epidemiological data, which revealed inflammation as a potential immuno-metabolic pathomechanism influencing blood mtDNA levels and mortality.

### 4.2. Limitations

We recognize certain limitations to our research. Although our sample size was comparable to similar studies, the smaller number of participants and death events in the follow-up phase limited the statistical power, especially for cause-specific analyses. Although we divided males and females in equal proportion into the blood mtDNA tertiles, this may not fully account for sex-related differences, as confounding factors, such as lifestyle, could impact sexes differently. Future studies are needed to better characterize these sex-specific effects. Additionally, we were able to conduct blood gene expression analysis only from the follow-up population, which prevented integrated analyses across both time points and limited the strength of the transcriptomic findings.

### 4.3. Impact and future perspectives

In our study, blood mtDNA abundance shows promise as a predictive biomarker for general morbidity and mortality up to three decades in advance. Preliminary evidence also suggests that decreases in blood mtDNA may be reversible [66]. Further investigations should explore interventions targeting the increase of blood mtDNA levels possibly via lifestyle changes as a means of primary prevention. Another direction could be the development of a diagnostic method for blood mtDNA abundance, as it potentially represents a sensitive and cheap laboratory test. While our findings also highlight limited specificity, many established clinical markers, such as C-reactive protein, similarly lack high specificity.

Future studies are needed to elucidate the pathomechanisms behind the decrease in blood mtDNA abundance in more detail. Importantly, reduced mitochondrial content and dysfunction should be considered alongside the effects of chronic low-grade inflammation, which can alter white blood cell composition and consequently influence the measured blood mtDNA levels. Based on our results, studying blood mtDNA content correlation with neutrophil counts and neutrophil-to-lymphocyte ratio in larger populations can yield valuable insights.

### 4.4 Conclusion

We demonstrated that the abundance of mtDNA in peripheral blood is associated with general morbidity and mortality over the span of three decades. We showed that blood mtDNA abundance lacks disease specificity and that the pathomechanism underlying its different levels may be related to subclinical inflammatory processes of the innate immune system, which are common across various etiologies. Given the relative simplicity of its quantification and the substantial follow-up time of our study, blood mtDNA abundance appears to be a sensitive and practical predictive biomarker for the primary prevention of diverse diseases in a middle-aged general population. These features make it a potential tool for reducing healthcare costs in the globally aging population.

## 5. Ethics statement

This study was approved by the Ethics Committee of the Medical Department of the University of Oulu (48/2009). All participants provided informed written consent, and they were informed of the opportunity to withdraw the consent at any time.

## 6. Data availability statement

Data contain sensitive patient information. The Regional Medical Research Ethics Committee of the Wellbeing services county of North Ostrobothnia, Finland has imposed these legal restrictions.

Contact information for ethics committee, to which data requests may be sent:

The Regional Medical Research Ethics Committee of the Wellbeing services county of North Ostrobothnia, Finland

Kajaanintie 50, 90220 Oulu

## 7. Author contributions

**Conceptualization**: Attila A. Sebe, Eija Pirinen, Olavi Ukkola.

**Methodology**: Attila A. Sebe, Juulia H. Lautaoja-Kivipelto, Jari Jokelainen, Juho T. Väänänen, Sini Skarp, Karri Parkkila, Risto Kerkelä, Eija Pirinen, Olavi Ukkola.

**Data curation**: Attila A. Sebe, Juulia H. Lautaoja-Kivipelto, Juho T. Väänänen.

**Formal analysis**: Attila A. Sebe, Juulia H. Lautaoja-Kivipelto, Jari Jokelainen, Juho T. Väänänen.

**Writing (original draft)**: Attila A. Sebe, Juulia H. Lautaoja-Kivipelto, Juho T. Väänänen.

**Writing (Review & Editing)**: Attila A. Sebe, Juulia H. Lautaoja-Kivipelto, Eija Pirinen.

**Visualization**: Attila A. Sebe, Juulia H. Lautaoja-Kivipelto.

**Software**: Jari Jokelainen.

**Project administration**: Eija Pirinen, Olavi Ukkola.

**Funding acquisition**: Eija Pirinen, Olavi Ukkola.

**Supervision**: Eija Pirinen, Olavi Ukkola.

## 8. Acknowledgements

We would like to thank Saija Kortetjärvi, University of Oulu, for valuable laboratory work; Maheswary Muniandy, University of Helsinki, for assisting in bioinformatics; and software developer Robert Osvath for support in processing raw laboratory measurement data. During the preparation of this manuscript, the authors used artificial intelligence tools solely to improve readability and grammar.

## 13. Supporting information captions

**S1 Figure. Blood mtDNA abundance demonstrated a sex-specific difference at baseline**. (A) At baseline, blood mtDNA abundance was higher in males than in females. (B) At follow-up, no sex difference was observed. Median value of each group is shown above the violin plots. In (A), *n*=505 in males and *n*=524 in females and in (B), *n*=281 in males and *n*=318 in females.

**S1 Table. Gene list used to calculate the OXPHOS score.**

Pathway information was acquired from the Ingenuity Pathways Analysis database. S1 Table is related to Fig 3.

**S2 Table. Gene list used to calculate the neutrophil degranulation (NeDegra) score.**

Pathway information was acquired from the Ingenuity Pathways Analysis database. S2 Table is related to Fig 3.

**S3 Table. Gene list used to calculate the mtDNA score.**

Pathway information was acquired from the Ingenuity Pathways Analysis database. S3 Table is related to Fig 3.

**S4 Table. Comparison of continuous variables across the male blood mtDNA tertiles in the baseline and follow-up populations.**

Data are presented as mean ± standard deviation (SD) or as median with 1^st^-3^rd^ interquartile range. Both SD and interquartile range are shown in brackets. *P*-values were calculated using one-way ANOVA followed by Tukey’s post hoc test or Kruskal-Wallis test followed by Mann-Whitney U-test. The male tertiles of low, moderate and high blood mtDNA abundance of the follow-up cohort are, by definition, not a fraction of the respective baseline tertiles. See Methods section for details.

**S5 Table. Comparison of continuous variables across the female blood mtDNA tertiles in the baseline and follow-up populations.**

Data are presented as mean ± standard deviation (SD) or as median with 1^st^-3^rd^ interquartile range. Both SD and interquartile range are shown in brackets. *P*-values were calculated using one-way ANOVA followed by Tukey’s post hoc test or Kruskal-Wallis test followed by Mann-Whitney U-test. The female tertiles of low, moderate and high blood mtDNA abundance of the follow-up cohort are, by definition, not a fraction of the respective baseline tertiles. See Methods section for details.

**S6 Table. Comparison of categorical variables across the male blood mtDNA tertiles in the baseline and follow-up populations.**

Data are presented as number of participants, with percentages shown in brackets for each mtDNA tertile. *P*-values were calculated using Pearson’s Chi-Square test or Fisher’s Exact Test (when the assumptions of the Pearson’s Chi-Square test were violated). The male tertiles of low, moderate and high blood mtDNA abundance of the follow-up cohort are, by definition, not a fraction of the respective baseline tertiles. See Methods section for details.

**S7 Table. Comparison of categorical variables across the female blood mtDNA tertiles in the baseline and follow-up populations.**

Data are presented as number of participants, with percentages shown in brackets for each mtDNA tertile. *P*-values were calculated using Pearson’s Chi-Square test or Fisher’s Exact Test (when the assumptions of the Pearson’s Chi-Square test were violated). The female tertiles of low, moderate and high blood mtDNA abundance of the follow-up cohort are, by definition, not a fraction of the respective baseline tertiles. See Methods section for details.

**S8 Table. Cox proportional hazards analyses results for all-cause mortality in the baseline and follow-up populations.**

At both time points, the final adjustment model and alternative adjustment model is shown. S8 Table is related to Fig 1.

CVD, cardiovascular diseases; HR, hazard ratio; CI, confidence interval.

**S9 Table. Cox proportional hazards analyses results for cause-specific mortality and cardiovascular morbidity in the baseline and follow-up populations.**

All analyses used the final adjustment model from the corresponding all-cause mortality analysis (baseline or follow-up). S9 Table is related to Fig 2.

HR, hazard ratio; CI, confidence interval.

**S10 Table. Significantly altered pathways according to IPA across blood mtDNA tertiles 1 (low) and 3 (high) in the follow-up population.**

Analysis was carried out using differentially expressed global genes at *p* < 0.05 and |log₂ fold change| > 0.2. Gene-level *p*-values used to define differential expression are distinct from the pathway *p*-values reported by IPA, which reflect statistical enrichment of genes mapping to each canonical pathway. All significantly altered pathways are shown (pathway *p*-value), irrespective of z-scores. IPA z-scores represent predicted activation state of the pathways, with positive observed z-scores indicating “activation” and negative z-scores indicating “inhibition”. Pathways with z-scores of “N/A”, for which IPA could not predict directionality, were excluded from further analyses and are shown at the bottom of the table. Remaining pathways are ranked by pathway *p*-values. The ratio represents the number of genes from our dataset that map to the pathway divided by the total number of genes in that canonical pathway in the IPA knowledge database. S10 Table is related to Fig 3.

**S11 Table. Significantly altered pathways according to IPA across blood mtDNA tertiles 2 (moderate) and 3 (high) in the follow-up population.**

Analysis was carried out using differentially expressed global genes at *p* < 0.05 and |log₂ fold change| > 0.2. Gene-level *p*-values used to define differential expression are distinct from the pathway *p*-values reported by IPA, which reflect statistical enrichment of genes mapping to each canonical pathway. All significantly altered pathways are shown (pathway *p*-value), irrespective of z-scores. IPA z-scores represent predicted activation state of the pathways, with positive observed z-scores indicating “activation” and negative z-scores indicating “inhibition”. Pathways with z-scores of “N/A”, for which IPA could not predict directionality, were excluded from further analyses and are shown at the bottom of the table. Remaining pathways are ranked by pathway *p*-values. The ratio represents the number of genes from our dataset that map to the pathway divided by the total number of genes in that canonical pathway in the IPA knowledge database. S11 Table is related to Fig 3.

**S12 Table. Significantly altered pathways according to IPA across blood mtDNA tertiles 1 (low) and 2 (moderate) in the follow-up population.**

Analysis was carried out using differentially expressed global genes at *p* < 0.05 and |log₂ fold change| 0.2. Gene-level *p*-values used to define differential expression are distinct from the pathway *p*-values reported by IPA, which reflect statistical enrichment of genes mapping to each canonical pathway. All significantly altered pathways are shown (pathway *p*-value), irrespective of z-scores. IPA z-scores represent predicted activation state of the pathways, with positive observed z-scores indicating “activation” and negative z-scores indicating “inhibition”. Pathways with z-scores of “N/A”, for which IPA could not predict directionality, were excluded from further analyses and are shown at the bottom of the table. Remaining pathways are ranked by pathway *p*-values. The ratio represents the number of genes from our dataset that map to the pathway divided by the total number of genes in that canonical pathway in the IPA knowledge database. S12 Table is related to Fig 3.

**S13 Table. Cox proportional hazards analyses results for all-cause mortality of the neutrophil degranulation (NeDegra) and mtDNA scores in the follow-up population.** Both analyses used the final adjustment model of the follow-up population. S13 Table is related to Fig 3.

