## Supplemental Figure 1 for "Lower mitochondrial DNA abundance in blood cells is associated with higher general morbidity and all-cause mortality: a 30-year prospective epidemiological study"

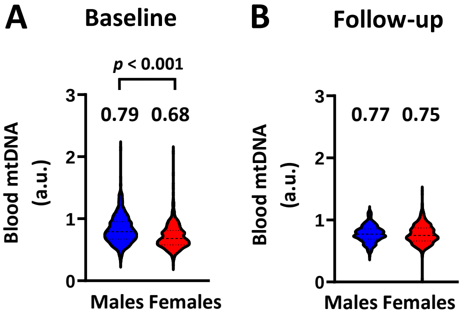


**S1 Figure. Blood mitochondrial DNA (mtDNA) abundance demonstrated a sex-specific difference at baseline**. (A) At baseline, blood mtDNA abundance was higher in males than in females. (B) At follow-up, no sex difference was observed. Median value of each group is shown above the violin plots. In (A), n=505 in males and n=524 in females and in (B), n=281 in males and n=318 in females.
